# Deep Sequence Modeling for Pressure Controlled Mechanical Ventilation

**DOI:** 10.1101/2022.03.02.22271790

**Authors:** Abdelghani Belgaid

## Abstract

This paper presents a deep neural network approach to simulate the pressure of a mechanical ventilator. The traditional mechanical ventilator has a control pressure monitored by a medical practitioner, which could behave inaccurately by missing the proper pressure. This paper exploits recent studies and provides a simulator based on a deep sequence model to predict the airway pressure in the respiratory circuit during the inspiratory phase of a breath given a time series of control parameters and lung attributes. This approach demonstrates the effectiveness of neural network-based controllers in tracking pressure waveforms significantly better than the current industry standard and provides insights to build effective and robust pressure-controlled mechanical ventilators.

## 1. Introduction

The mechanical ventilator is a critical medical component in an intensive health environment, especially during the COVID-19 pandemic. The current industry standard of PID controllers uses a combination of proportional, integral, and derivative controls that takes the deviation from the target waveform as input and adjusts it in the pressure chamber to correct the output waveform and reduce the deviation gap. Consequently, the PID controller relies on medical practitioners’ continuous manual monitoring and adjustment, leading to incorrect pressure. Although it satisfies the needs and safety requirements, it cannot generalize and adapt quickly to different clinical conditions. Thus, a dynamic controller that constantly adjusts its pressure might solve this problem. That is where machine learning ingress.

As it is known, machine learning models are data-hungry and require a substantial quantity of data that is not easy to obtain. Therefore, two types of machine learning models are required in this case: the first model represents a simulator that generates data which will enable the second one to be fine-tuned using this data and replace the traditional PID controller algorithm.

A recent paper demonstrated the potential advantage of machine learning for ventilator control (Suo et al., 2021) on an open-source ventilator (LaChance et al., 2020) designed in response to the COVID-19 pandemic, paving the way for intelligent control methods that are robust and require less manual monitoring. We exploit this study and present a simulator based on a deep sequence model and leverage statistical learning techniques to improve the model’s enhancement.

Google Brain and Princeton University released the dataset on a Kaggle competition (Google Brain, 2021), consisting of approximately 125,000 simulated breaths, 60% of which were dedicated to the training phase. Each breath is a sequence of 80 timesteps that includes six features. We will use this dataset to train a stack of Residual Bidirectional Long Short Term Memory or ResBiLSTM to predict such a sequence during the inspiratory phase for each breath and measure the score with the mean absolute error (MAE) metric.

Although the ‘no free lunch theorem’ is applied, we will find additional methods to predict more accurate ventilator pressure values. Our contribution will not be limited to the model-based ventilator simulator. However, it will also provide a feature and statistical perspective to counter the challenges that arise, particularly the problem of generalizing in a dynamical system across lungs with varying physical characteristics.

## 2. Background

The challenges of the traditional mechanical ventilator with PID controls described previously include continuous manual monitoring and adjustment by a medical practitioner, missing the proper target pressure, and costly development of new methods of mechanical ventilators, even before reaching clinical trials, leading to the development of simulators to overcome these challenges.

The recent COVID-19 pandemic has given rise to a significant number of publications of open-source ventilator designs and opened the way for several temptations to reinvent these ventilators by introducing intelligent systems and techniques that employ decision support approaches based on machine learning models tuned to accommodate the patient response to improve the accuracy of their pressure values.

Machine Learning for Mechanical Ventilation Control (Suo et al., 2021) is the most significant one. The paper presents two models: the real2sim creates a ventilator simulator based on deep feedforward networks and the sim2real where the data generated by this simulation is used to tune a controller. Our paper aims to improve real2sim by introducing a deep sequence model with stacked Long Short-Term Memory (Hochreiter and Schmidhuber, 1997) combined with other advanced deep learning methods and techniques (He et al., 2015; Schuster and Paliwal, 1997).

## 3. Methodology

### Data Description

The dataset was collected from a modified open-source ventilator, designed by the People’s Ventilator Project (PVP) at Princeton University (LaChance et al., 2020), and connected to an artificial bellows test lung (IngMar, 2020) via a respiratory circuit (Suo et al., 2021). Figure 1 below illustrates the circuit, with the two control inputs highlighted in green and the state variable (airway pressure) to predict in blue.

**Figure 1.**
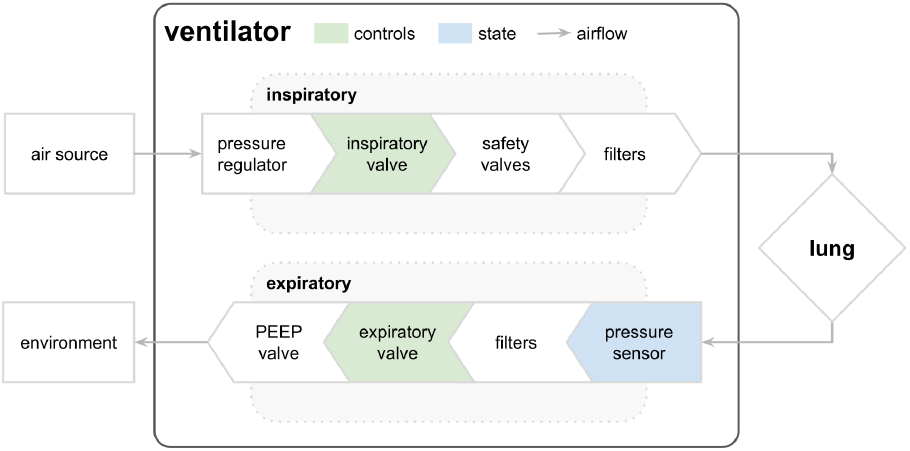
A simplified respiratory circuit shows the airflow through the inspiratory pathway, entering and exiting the lung, and exiting through the expiratory pathway. (Suo et al., 2021)

The dataset has eight features with approximately 75,000 training breaths and 50,000 test breaths (with over six million rows). It contains details regarding lung attributes, the input and output valves of the machinery, and the pressure inside the lungs connected to the machine.

The key attributes are the control `u` and `pressure` over time and a range of lung attributes (`R`, `C`). Each time series represents a breath of approximately three-second, and each breath is composed of an inspiratory phase and an expiratory phase. The dataset is organized such that each row is a timestep in a breath and yields the two control signals, the resulting airway pressure, and relevant attributes of the lung.

The first control input `u_in` is a continuous variable from 0 to 100, representing the percentage of the inspiratory solenoid valve is open to let air into the lung (i.e., 0 is completely closed and 100 is completely open). The second control input `u_out` is a binary variable representing whether the exploratory valve is open (1) or closed (0) to let the air out.

During the inspiratory phase, the target applied pressure increases to the maximum inspiratory pressure (PIP). The target decreases to the positive end-expiratory pressure (PEEP) during the expiratory phase, maintained to prevent the lungs from collapsing. The PIP and PEEP values, along with the duration of these phases, define the time-varying target waveform, which is specified by a medical practitioner. The ventilator’s task is to ensure the target waveform is replicated for different lungs, thus requiring a device to handle these differences. There are two main discrete parameters that describe the differences between lungs. The first parameter is resistance `R` which represents the change of pressure per variation in flow (air volume per time) in cmH2O/L/S. The second parameter is compliance `C,` which is the capacity of the lung to expand, representing the change in volume per change in pressure in mL/cmH2O. In the case of the Covid-19, the values of `R` &`C` most often found in patients were `R` equal to 20 and `C` among these values [10, 20, 50] (Haudebourg et al., 2020).

### Data Processing

- **Feature Engineering**

The features consist of 88 features, which can be divided into 3 original features [`time_step`, `u_in`, `u_out`] and 85 engineered features. For the original features, all features except `R` and `C` were used in their primary forms, and one-hot encoded `R` and `C` were used to show the type information of `R` and `C` to the model. After performing exploratory data analysis, we designed more than 70 engineered features that utilize the available features, converge the model faster, and reach a low MAE score. In addition, multiple statistical data quality checks, such as multicollinearity and serial correlation, were performed without noticeable results or even harming the model. Many of the features have been discussed in the competition forum. Additional details on these features are given in the GitHub repository^1^.

- **Data Normalization**

The dataset is scaled since it increases the learning speed of the neural network, reduces the risk of numerical overflow, and limits the influence of outliers. RobustScaler is used as a data scaling technique to normalize the datasets with a more comprehensive quantile range of [20,80] to reduce variance. RobustScaler is robust to outliers since adding or removing outliers in the training set will yield approximately the same transformation. As a result, outliers do not significantly influence it. Moreover, it will not affect the one-hot encoded features since the one-hot encoded variables [`R`, `C`, `R`_`C]`have a median of 0 and an interquartile range of 0 or 1.

### Model Architecture

The model is based on stacked residual bidirectional LSTM (ResBiLSTM) with 4 bidirectional LSTM layers with [1024, 512, 256, 128] units, and 2 final dense layers with [128, 1] units (Figure 2). The penultimate layer is employed with scaled exponential linear units (SELU) activation. The ResBiLSTM architecture extracts the semantic information in the paths with specific patterns. The LSMT layers are necessary since the target pressure is heavily reliant and dependent on previous time points. Additionally, we add skip connections by feeding the *l-th* layer output as the *l+2-th* layer input, thus avoiding performance degradation. We use the SELU as an activation in the penultimate layer since it addresses the vanishing gradient problem and helps train deep networks with many layers, employs strong regularization, and makes learning highly robust even in the presence of noise and perturbations (Klambauer et al., 2017).

**Figure 2.**
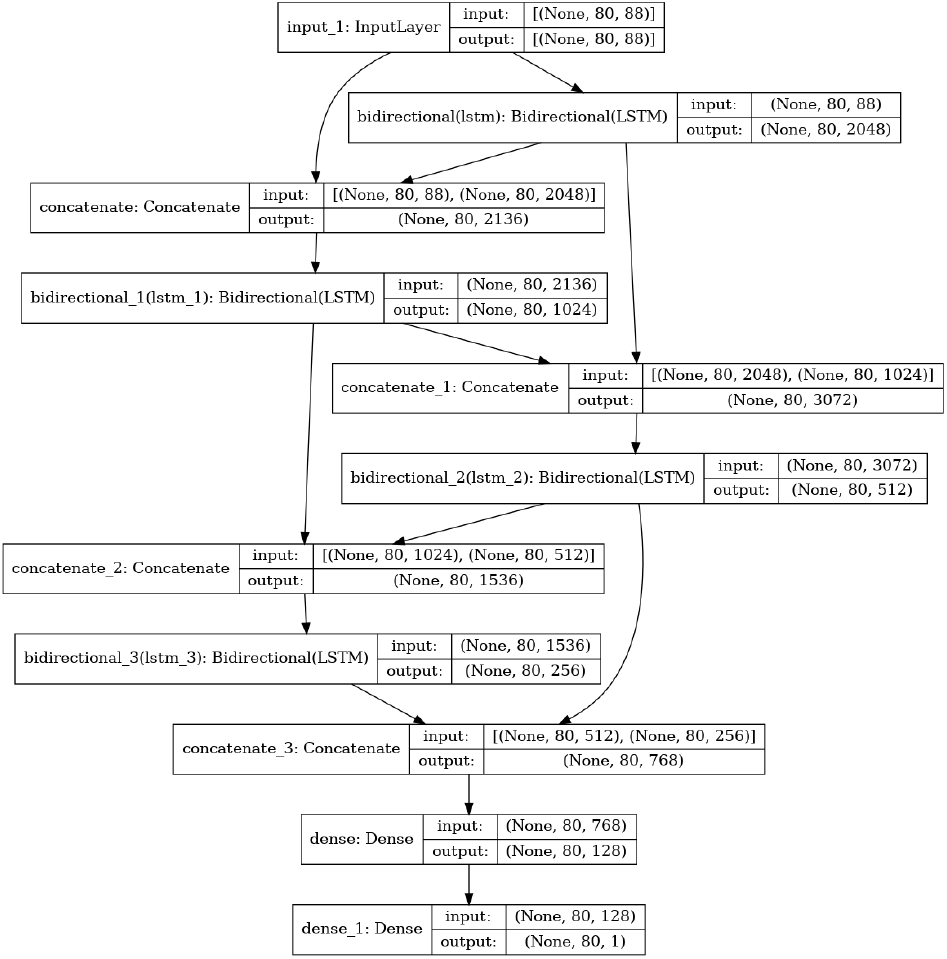
Model Architecture based on ResBiLSTM

### Experimental Details

- **Training Details**

The solution is implemented with Scikit-learn and TensorFlow to process the data and build the model. The model was trained on approximately 60% of the dataset with 200 epochs, five multiple folds, and a batch size of 512. The training was on a TPU v3.8 and took nearly 4 hours to train the models. The model’s initial hyperparameters are mostly in default state or taken for the original papers. We use Adam as an optimizer and ReduceLROnPlateau as a learning rate scheduler. We determine when to stop training with an early stopping based on the MAE score on the validation set. Due to the limited computation power, we implement the learning rate scheduler with 5 patience steps and 15 patience steps on early stopping. Although dropout is an effective way to regularize a model, it was surprisingly ineffective in early training trials whence it was not implemented. For the validation strategy, we will perform five-fold cross-validation, and the results go through the post-processing methods described below. Finally, we evaluate the results according to the mean absolute error.

- **Metric**

The score is measured with the mean absolute error (MAE) between the predicted and actual pressures during the inspiratory phase of each breath. The expiratory phase is not scored. The score is given by:

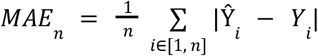

where *n* ∈ ℝ, **(Y, Ŷ)** ∈ ℝ^(80×1×2)^, and where Ŷ is the vector of predicted pressures and **Y** is the vector of actual pressures.

- **Post-processing**

The post-processing techniques consist of five methods to compute the ensemble folds results: mean, median, median with round prediction, mean-median method with spread a limit of 50, and rounding predictions to the nearest neighbors. The mean-median method^2^ consists of a combination of the mean and the median, where it returns the mean of a prediction if it is not an outlier or the median otherwise. The rounding prediction method with median returns the nearest pressure value in the training set, which is only 950 discrete possible values.

## 4. Experimental Results &Discussions

### Quantitative Results

The model described above achieved over the five folds cross-validation an MAE score of 0.15 on the holdout dataset (Figure 3). Table 1 summarizes the MAE scores on the testing set by each post-processing method. The main challenge in training DNN is to avoid overfitting. Therefore, various methods were implemented, such as dropout and label smoothing, but conduct no obvious or even worsens the results.

**Table 1.** MAE scores by method over five folds

| Method | Ensemble |
| --- | --- |
| Mean | 0.1361 |
| Median | 0.1347 |
| Mean-Median | 0.1338 |
| Median Rounded | 0.1335 |
| Nearest Neighbor | **0.1322** |

**Figure 3.**
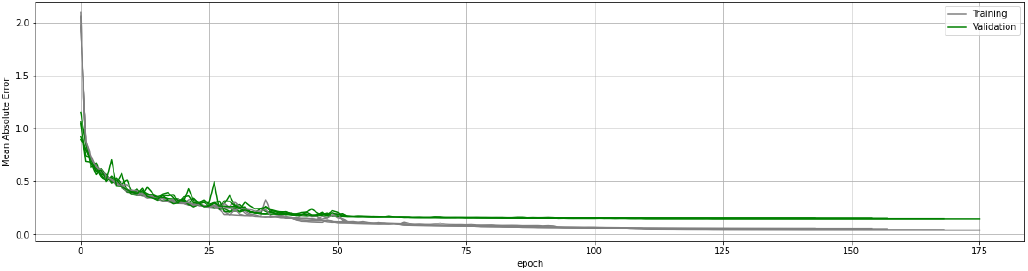
Evolution of training & validation scores

### Further Improvements

Our proposed solution might be over-engineered, thus reducing and skipping some computations might improve the score and the model explainability. The following is a list of enhancements that may improve simulator accuracy:

- Switch from a regression task to a classification one. The method is feasible since there are only 950 possible output values in the training set;
- Perform data augmentation, e.g., masking augmentation randomly performed on the `R` and `C` only, random shuffling of units within a specific short window in a sequence, and mixup augmentation in which two sequences are randomly selected and mixed. Other augmentations such as Cutout and CutMix can also be implemented;
- Perform Fourier or Wavelets transforms to eliminate noise, create approximations of different values movements and generalize short and long-term trends;
- Overparameterization of neural networks (Dar et al., 2021; Power et al., 2021), especially with a large number of epochs and heavy dropout rate, might be beneficial for long term fitting;
- Multitask LSTM: adjust the model to predict secondary targets, thus adding additional targets to the original one to reduce overfitting and improve generalization. New targets such as pressure lags, cumulative pressure, and pressure variance at each timestep. These targets force the model to learn more about the main pressure target, its derivative, and its integral, thereby improving its performance and accelerating the convergence;
- Hybrid, multi-model, and adversarial networks that combine CNN, Transformers, and other RNN architecture.

## 5. Conclusion

This paper presented a simulator based on a ResBiLSTM to predict ventilator pressure and generate future data. Additionally, we highlighted potential refinements on the model, improvements to the dataset, and identified other potential architectures. This solution still needs more detailed consideration before any clinical trial. Nevertheless, we believe that the insights in this paper will help healthcare researchers build effective and robust pressure-controlled mechanical ventilators with intelligent systems.

## Data Availability

All data produced in the present work are contained in the manuscript.

## Acknowledgments

I would like to express my profound appreciation to the Kaggle community for sharing their notebooks and discussions, which inspired this work. I would also like to thank Google Brain, Princeton University, and Kaggle for making the relevant materials available and hosting the competition.

## Footnotes

1 https://github.com/abdelghanibelgaid/nnvpp

2 https://www.kaggle.com/c/ventilator-pressure-prediction/discussion/282735

## References

1. Dar, Y., Muthukumar, V., & Baraniuk, R.G. (2021). A Farewell to the Bias-Variance Tradeoff? An Overview of the Theory of Overparameterized Machine Learning. arXiv preprint 2109.02355

2. Google Brain & Princeton University. (2021). Ventilator Pressure Prediction. Retrieved November 04, 2021, from https://www.kaggle.com/c/ventilator-pressure-prediction

3. Haudebourg, A.-F., Perier, F., Tuffet, S., De Prost, N., Razazi, K., Mekontso Dessap, A., & Carteaux, G. (2020). Respiratory Mechanics of COVID-19-versus Non-COVID-19-associated Acute Respiratory Distress Syndrome. American Journal of Respiratory and Critical Care Medicine, 202(2):287–290

4. He, K., Zhang, X., Ren, S., & Sun, J. (2015). Deep residual learning for image recognition. arXiv preprint 1512.03385

5. Hochreiter S., & Schmidhuber J. (1997). Long short-term memory. Neural Computation, 9(8):173580.

6. IngMar. Quicklung products. (2020). URL https://www.ingmarmed.com/product/quicklung/.

7. Klambauer, G., Unterthiner, T., Mayr, A., & Hochreiter, S. (2017). Self-Normalizing Neural Networks. arXiv preprint 1706.02515

8. LaChance, J., Zajdel, T. J., Schottdorf, M., Saunders, J. L., Dvali, S., Marshall, C., Seirup, L., Notterman, D. A., & Cohen, D. J. (2020). PVP1–The People’s Ventilator Project: A fully open, low-cost, pressure-controlled ventilator. https://doi.org/10.1101/2020.10.02.20206037

9. Power, A., Burda, Y., Edwards, H., Babuschkin, I., & Misra, V. (2021). Grokking: Generalization Beyond Overfitting on Small Algorithmic Datasets. arXiv preprint 2201.02177

10. Schuster, M. and Paliwal, K. (1997). Bidirectional recurrent neural networks. IEEE Transactions on Signal Processing, 45(11), 2673–2681.

11. Suo, D., Zhang, C., Gradu, P., Ghai, U., Chen, X., Minasyan, E., Agarwal, N., Singh, K., LaChance, J., Zajdel, T., Schottdorf, M., Cohen, D., & Hazan, E. (2021). Machine Learning for Mechanical Ventilation Control. arXiv preprint 2102.06779

